# Implementation and Preliminary Evaluation of a Therapeutic Communication Educational Module for Nursing Trainees in a Low-Resource Setting

**DOI:** 10.64898/2026.02.19.26346685

**Authors:** Helen Mukakarisa, Aloysius Gonzaga Mubuuke, Rose Clarke Nanyoga, Patience A. Muwanguzi

**Author notes:** Corresponding author Helen Mukakarisa (HM).

## Abstract

**Introduction:** Therapeutic communication is the basis of nursing care yet it has been considered so stressful by student nurses with only 3.5% of nursing students in Kampala Uganda exhibiting optimum therapeutic communication competence. This has been attributed to inadequate training. Faculty must explore means to impart these skills in nursing students. This study implemented and evaluated an educational intervention module on therapeutic communication skills for nurses for incorporation into their teaching learning activities.

**Methods:** A one-group pre–post quasi-experimental study was conducted with 41 diploma extension nursing students, selected via census sampling. Data were collected using self-administered structured questionnaires (content validity = 0.98; Cronbach’s α = 0.96), on students’ knowledge and perceived confidence in performing therapeutic communication. Observation checklists were used to evaluate students’ ability to establish nurse–patient relationships and deliver bad news in the skills lab, both before and after the intervention.

**Results:** There was a significant improvement of knowledge scores from 4 (IQR: 3, 5) to 8.0 (IQR: 7.0, 9.0), (P value <0.001); perceived confidence in practicing therapeutic communication scores from 144.0 (IQR: 136.0, 153.0) to 164.0 (IQR: 155.0, 174.0) (P value <0.001); ability to initiate a nurse-patient relationship from 12.0 (IQR: 10.0, 14.0) to 17.0 (15.0, 18.0) (P value <0.001); and the ability to break bad news to the patient/caretaker from 9.0 (IQR: 7.0, 12.0) to 16.0 (14.0, 18.0) (P value <0.001) after the intervention. All scores improved in all categories of sex, program and semester of study for all participants apart from participants in the first semester of study.

**Conclusion:** This study offers preliminary evidence that the educational intervention improves nursing students’ therapeutic communication skills. Further longitudinal research is needed to assess the sustained effectiveness of the module, the teaching methods used, and patients’ perspectives on students’ TC competence.

## Introduction

Communication is widely recognized as the foundational medium through which health professionals deliver effective messages and facilitate meaningful change in patients’ health outcomes(1). Therapeutic communication (TC) between nurses and patients is an intentional interaction that occurs throughout the process of identifying and addressing patients’ health problems (2). Effective communication is particularly important because it directly contributes to the quality of patient care (3). Therapeutic communication is holistic and patient-centered, encompassing physiological, psychological, environmental, and spiritual dimensions of care (4–5). It also serves as a critical tool in supporting patients to express and manage emotional distress, thereby reducing anxiety and promoting emotional relief (6). To achieve positive patient outcomes, nurses must possess strong communication skills; consequently, communication and interpersonal relations should be deliberately integrated into nursing education through dedicated coursework and clinical training experiences (7).If students must establish therapeutic relationships with their patients and as future nursing professionals, they should thus be trained to be effective communicators(2). TC between the nurse and patient is considered one of the most significant clinical methods of communication and the basis of nursing care (8–9), yet has been considered to be so stressful by student nurses (3). Previous literature has indicated that student nurses had deficiencies in applying TC techniques(5). A recent study determined that only 3.5% of the Diploma Nursing Extension (DNE) students in Uganda could optimally communicate therapeutically yet the benefits of therapeutic communication are two fold for both the students and the clients. Inadequate communication between nurses and other health workers may also cause sentinel incidents in health services. This also contributes to job dissatisfaction, which tends to affect the quality of care and patient safety(6). These insights emphasize the urgent necessity for healthcare professionals to prioritize effective communication strategies to enhance patient care outcomes and overall quality of service delivery (6).

Nurses’ lack of effective communication skills has been related to inadequate training emphasizing the need for nurses to develop their communication skills and to receive adequate training from nursing faculty (7). For this reason, nurse faculty must explore means to impart therapeutic communication skills during training (8). Currently in Uganda, training in communication skills occurs during clinical rotations and is not structured to effectively equip students with these skills. A number of interventions have been implemented and have been found to be effective in improving Nurses TC. For example, the Nurse-led education interventions to improve nursing knowledge and understanding of the Situation, Background, Assessment and Recommendation (SBAR) tool used to exchange critical patient information concisely and efficiently (9), training sessions, comprehensive guides, specialized programs, and brief workshops(10). By prioritizing the development of communication skills, hospitals can improve patient care outcomes while also supporting the well-being of nursing staff (6). Given the central role of communication in nursing practice, investing in communication skills training represents a strategic priority for hospitals and training institutions. Such training not only equips nurses with essential competencies but also enhances their motivation and capacity to carry out professional roles and responsibilities effectively. Accordingly, this study aimed to pilot the implementation and evaluate the preliminary effectiveness of a therapeutic communication educational module for DNE students in Kampala, Uganda.

## Methods

### Study design

This study employed a single-group, pre–post quasi-experimental design. The TC interventional module was implemented and evaluated using a one-group pre–post approach to assess the effectiveness of the educational intervention on DNE students’ TC knowledge, perceived confidence in performing TC, and practical TC skills. The ADDIE instructional design model—Analysis, Design, Development, Implementation, and Evaluation—served as the overarching framework guiding the module’s development, implementation, and evaluation. The ADDIE model provides a systematic and rigorous approach to designing training materials aligned with evolving pedagogical priorities and has been widely applied in educational contexts (15). As this study aimed to pilot the TC module and examine its preliminary effectiveness, primary emphasis was placed on the Implementation and Evaluation phases of the ADDIE model, while the Analysis, Design, and Development phases informed the creation of the intervention. The figure below illustrates the steps of the ADDIE model and how the pre-posttest quasi-experimental study design was integrated into the implementation and evaluation steps of the model which was the focus of this particular study.

**Figure 1:**
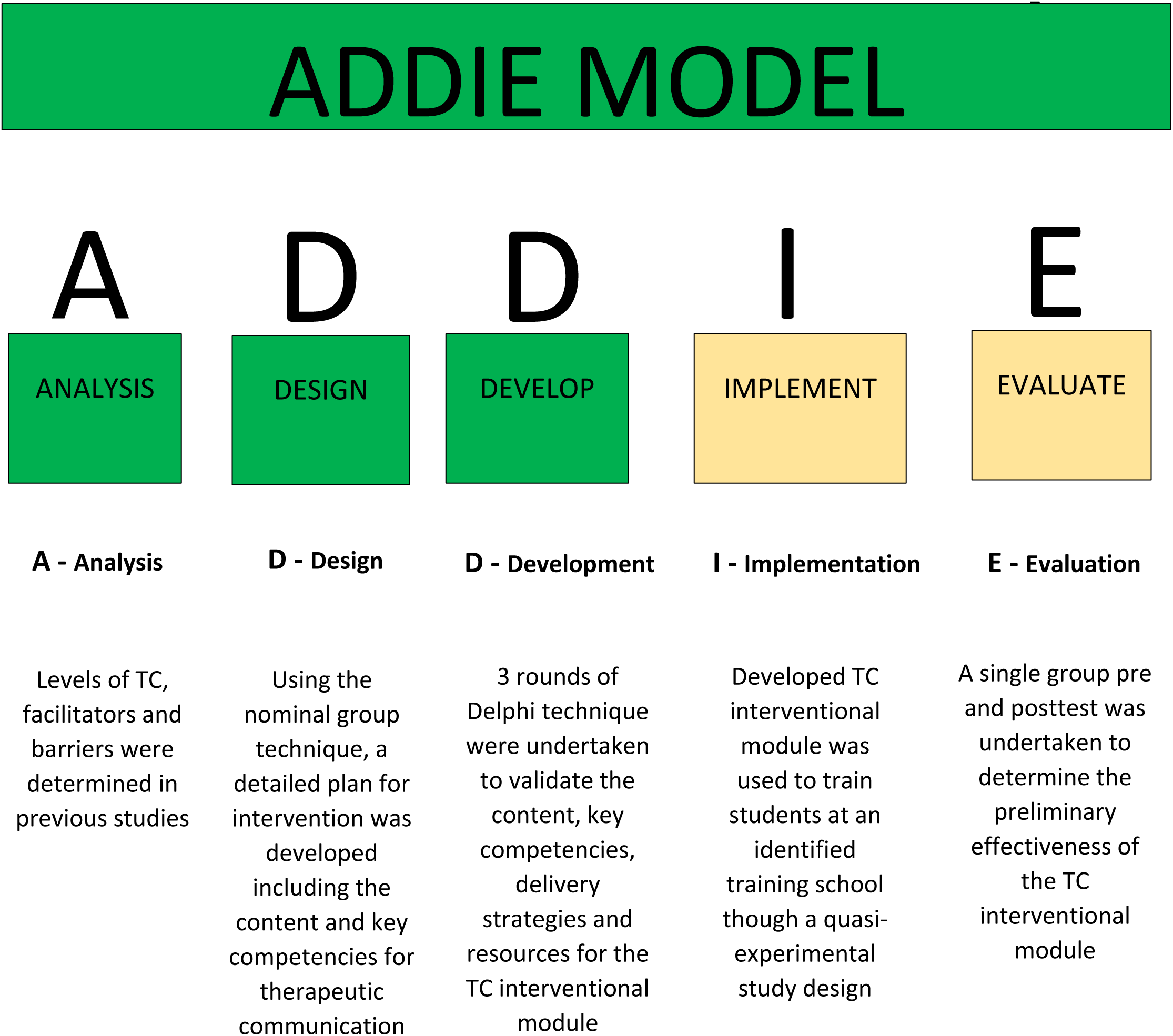
ADDIE Model.

Quasi experimental design is a research approach employed to estimate the causal impact of an intervention on a target population without randomization. In educational settings, quasi-experimental designs are the most suitable because total randomization and total control of variables are difficult to achieve but also because it is unethical to deny a willing learner a beneficial educational intervention (11).

### Setting and participants

This study was conducted at St. Francis School of Nursing and Midwifery (SoNM), Nsambya Hospital, a nursing training institution that offers diploma-level programs for individuals advancing from certificate-level qualifications. The study involved diploma nursing students enrolled in the Diploma in Nursing Extension program. In Uganda, Diploma Nursing Extensors—also known as post-certification trainees—are qualified nurses or midwives who hold a certificate in nursing or midwifery, have a minimum of two years of clinical work experience, and return to training to complete an 18-month post-certificate diploma program. St. Francis SoNM is a faith-based institution affiliated with the Catholic Church with a total enrollment of 690 students across certificate and diploma nursing and midwifery programs. The pilot site was purposively selected. Participants were eligible for inclusion if they had at least two years of post-certification clinical experience and provided written informed consent. Students who were absent from school for any reason during the data collection period were excluded from the study.

### Sample size estimation and sampling procedure

This pilot study adopted recommendation for pilot studies for continuous outcomes that a sample size ≥35 is optimal for assessment of preliminary effectiveness of an intervention (12). This pilot enrolled 41 students of diploma extension in nursing and midwifery at St. Francis-Nsambya SoNM. Who were selected using census sampling technique, where all eligible participants available were enrolled in the study.

### Data collection tools and procedure

Study participants were recruited from the training school in November 2024. Those who provided informed consent were screened for eligibility and subsequently completed a pretested, self-administered structured questionnaire. The questionnaire captured participants’ demographic characteristics, perceived confidence in performing therapeutic communication (TC), and knowledge of TC. Data collection was conducted using a structured instrument comprising three sections: demographic information, perceived confidence in performing TC (49 items), and TC knowledge assessed through 10 multiple-choice questions.

The questionnaires were self-administered by participants both before and after the educational intervention, with guidance provided by trained research assistants. In addition, trained assessors used observation checklists to evaluate participants’ practical TC skills, specifically their ability to establish a nurse–patient relationship and to deliver bad news. These assessments were conducted in the skills laboratory and were performed once before and once after the intervention.

The perceived confidence in performing TC scale (49 items) had been previously validated among student nurses in Uganda, demonstrating good psychometric properties, including a convergent validity of 0.74, divergent validity of −0.19, a scale content validity index of 0.98, and high internal consistency reliability (Cronbach’s alpha = 0.96).

### Study variables

The primary outcomes for the pilot study was knowledge on TC, perceived confidence in performing TC, and the ability to perform TC in a skills lab (specifically ability to initiate a nurse/midwife-patient relationship and the ability to break bad news to a patient). The participants’ perceived confidence in practicing TC was assessed using TCC questionnaire that was previously validated for student nurses in Uganda. The independent variables included the age, sex, program of study, semester of study, and years of working experience prior to enrolling for the diploma extension program.

### Data management

The data was collected through hardcopy questionnaires. The data was later entered into data entry screens designed in Epidata software. The entry screens were designed with logic commands to ensure accurate data entry

### Data analysis

The knowledge about TC was assessed with a set of 10-multiple choice questions for which a correct answer was weighted “1” and otherwise “0”. For the perception of TCC 49 4-level Likert item questions weighted “1” to “4” where a higher score represents more confidence in performing TC. A total score was computed before and after the intervention. The sum of the score was computed to get the total knowledge and perception scores before and after an intervention. As far as practice of TC is concerned, the ability to create a nurse-patient relationship was assessed by observing the participants while following the important phases of initiating a nurse-patient relationship including pre-interaction, orientation, working and termination phase. The steps were evaluated by a trained nurse on a scale of 0 to 20 where a higher score represented greater ability to create a nurse-patient relationship. The ability to break bad news was assessed by observing the participants following a stepwise approach of breaking bad news. The trained nurse evaluated the students as they performed the procedure on a scale of “0” to “20” where a higher score represents greater ability to break bad news.

Data was analyzed using STATA version 17.0 (Texas, USA). The participant characteristics were summarized using descriptive statistics where continuous variables were summarized using the median and interquartile range (IQR), and categorical variables were summarized using the frequencies and percentages. The primary outcomes were knowledge on therapeutic communication, perceived confidence in performing therapeutic communication, ability to initiate a nurse-patient relationship and the ability to break bad news to the patient/caretaker. All the outcomes were continuous outcomes and were summarized using the median and IQR. The outcomes were compared before and after the educational intervention using the Wilcoxon-signed rank test with the level of significance set at <0.05. Sub-group analysis of the primary outcomes across demographic characteristics of sex, program and semester of study before and after the intervention was done and level of significance set at <0.05.

### Quality control

The research assistants who collected data were nurses or midwives with at least a Bachelor’s degree in nursing or midwifery, and were further trained on the protocol and study procedures. The questionnaires were pre-tested before use to ensure that they were understandable to the participants. The questionnaires were assessed for completeness before the participants left the site to ensure that all the necessary data was captured.

### Ethical considerations

Ethical approval to conduct the study was obtained from the Makerere University School of Medicine Research Ethics Committee (Mak-SOMREC-2022-341), and the Uganda National Council of Science and Technology (HS3180ES). The administrative clearance was obtained from the head of the nursing school. All the participants enrolled in the study provided written informed consent prior to enrollment, after receiving information about the study following required local and international ethical guidelines. The study materials were kept under lockable cabinets only accessible by the study team, and all the dissemination information sheets and publications were de-identified to preserve confidentiality.

### Description of the Therapeutic communication intervention that was implemented

The therapeutic communication (TC) module was developed following the needs analysis, design, and development phases of the ADDIE framework. Using the Nominal Group Technique and validated through the Delphi method, the final module comprised five topics and 22 subtopics, totaling 45 hours. It aims to equip students with verbal and non-verbal communication skills to convey respect and empathy, encourage patient expression, and maintain professional boundaries. The module covers the basics of TC, theoretical foundations, the process of building therapeutic relationships, and application of various TC techniques. By the end of the training, students are expected to create environments conducive to effective communication, demonstrate professional attitudes, and foster trust in interactions with patients across diverse backgrounds and situations. The module was delivered using a blended learning approach, combining face-to-face classroom sessions with online learning. Active teaching strategies, including brainstorming, small group discussions, role play, simulations, demonstrations and return demonstrations in the clinical skills laboratory, and guided observations of therapeutic communication in clinical settings, were employed. Details are found in the S1table attached.

Competence was defined as the ability to perform tasks expected in the workplace. To achieve this, students were required to attend at least 75% of the course and complete both continuous assessments and end-of-semester examinations. Continuous assessment accounted for 40% of the overall grade and included both theoretical and practical components. Theoretical assessments comprised assignments (class exercises and take-home assignments, 10%), individual class tests (10%), and reflective journals (20%). Practical assessments included skills laboratory work, practical tests or projects (10%), clinical assessments (10%), and completion of a clinical logbook (20%), in accordance with the national DNE curriculum.

### Implementation of the TC interventional module

In the ADDIE model, the implementation phase is a crucial stage for testing and refining educational content in a real-world setting (18). During the implementation of the TC module, students were first provided with a brief overview of the rationale for developing the intervention. They then participated in 45 hours of training delivered through both face-to-face and virtual sessions, which included classroom instruction, clinical skills laboratory practice, and clinical area exposure. Active teaching strategies—such as brainstorming, group discussions, simulations, and role plays—were used in classroom sessions, while demonstrations and return demonstrations were conducted in the skills laboratory. In clinical areas, guided observations allowed students to watch qualified staff apply therapeutic communication with patients. At the end of each session, students were given scenario-based assignments for self-reflection.

## Results

### Demographic characteristics of study participants

Most of the participants were female (87.8%) with a median age of 27 years (IQR: 26, 28).Majority were in the Diploma in Nursing program (73.2%) and were in semester three of study (73.2%). The participants had worked for a median of 3 years (IQR: 3, 4) prior to enrolling for a diploma extension program. The characteristics of participants are summarized in Table2 below.

**Table 2:**
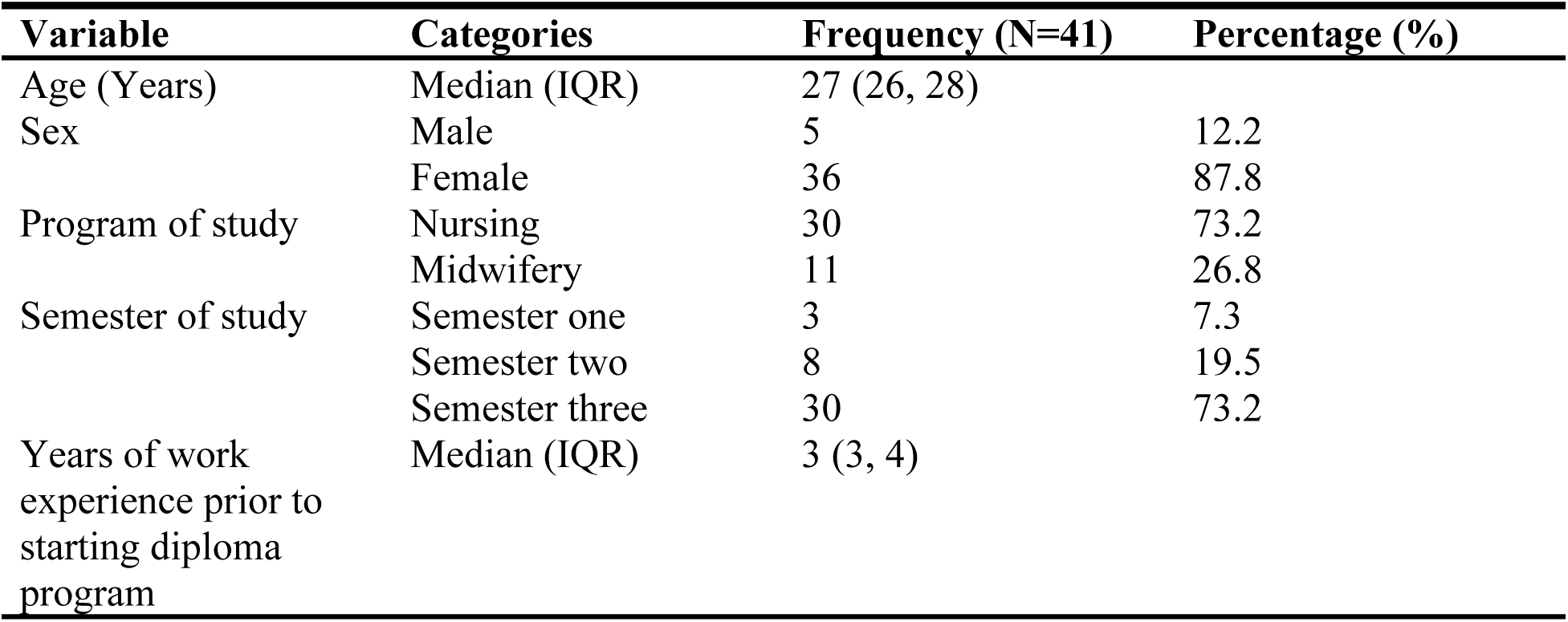
Characteristics of 41 students that participated in the study.

### Participants’ Knowledge about TC

The median knowledge scores of the participants significantly improved from 4 (IQR: 3, 5) at baseline to 8.0 (IQR: 7.0, 9.0) after an educational intervention (P value <0.001). The knowledge scores improved in all categories of sex and program (P values <0.05), however, no significant improvement in knowledge scores was observed among participants in semester one of study (P value 0.109) as is represented in Table3.

**Table 3:**
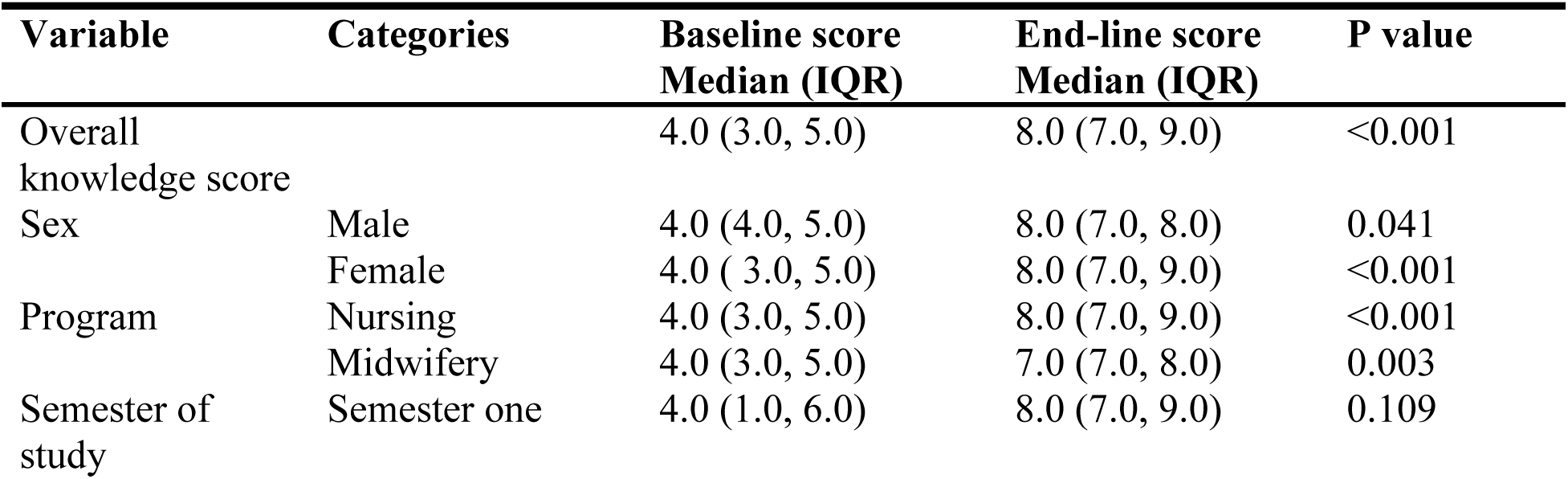
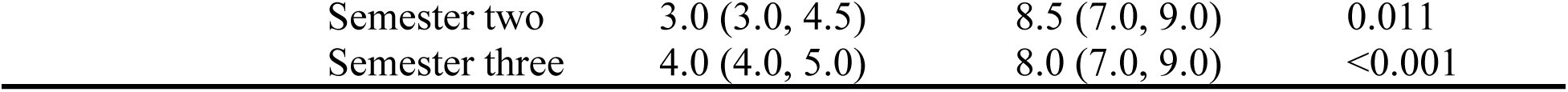
Knowledge scores before and after an educational intervention among students who participated in the study.

### Participants’ perceived confidence in performing TC

The participants’ perceived confidence in practicing therapeutic communication scores significantly improved from baseline median of 144.0 (IQR: 136.0, 153.0) to 164.0 (IQR: 155.0, 174.0) after the intervention (P value <0.001). The participants’ confidence in practicing therapeutic communication scores improved in all categories of sex, program and semester of study (P values <0.05) apart from participants who were in the first semester of study (P value 0.103). The findings are presented in Table 4 below.

**Table 4:** Participants’ perceived confidence in performing TC scores before and after an educational intervention.

| Variable | Categories | Baseline score<br>Median (IQR) | End-line score<br>Median (IQR) | P value |
| --- | --- | --- | --- | --- |
| Overall confidence in performing TC score |  | 144.0 (136.0, 153.0) | 164.0 (155.0, 174.0) | <0.001 |
| Sex | Male | 136.0 (126.0, 146.0) | 156.0 (148.0, 171.0) | 0.042 |
|  | Female | 144.5 (138.0, 154.5) | 165.0 (155.5, 174.0) | <0.001 |
| Program | Nursing | 143.0 (128.0, 153.0) | 166.5 (155.0, 174.0) | <0.001 |
|  | Midwifery | 146.0 (141.0, 155.0) | 163.0 (155.0, 174.0) | 0.004 |
| Semester of study | Semester one | 143.0 (136.0, 159.0) | 155.0 (148.0, 177.0) | 0.103 |
|  | Semester two | 141.0 (131.0, 153.5) | 171.0 (155.0, 176.0) | 0.012 |
|  | Semester three | 145.0 (137.0, 153.0) | 163.5 (156.0, 171.0) | <0.001 |

### Participants’ ability to initiate/create a nurse-patient relationship

The ability to initiate a nurse-patient relationship scores improved from a baseline median of 12.0 (IQR: 10.0, 14.0) to 17.0 (15.0, 18.0) after the intervention (P value <0.001). The ability to initiate a nurse-patient relationship scores improved in all categories of sex, program and semester of study (P values <0.05) apart from participants who were in the first semester of study (P value 0.109). This is presented in Table 5 below.

**Table 5:** Participants’ scores of their ability to initiate nurse-patient relationships before and after the educational intervention.

| Variable | Categories | Baseline score<br>Median (IQR) | End-line score<br>Median (IQR) | P value |
| --- | --- | --- | --- | --- |
| Overall ability to initiate nurse-patient relationship |  | 12.0 (10.0, 14.0) | 17.0 (15.0, 18.0) | <0.001 |
| Sex | Male | 10.0 (8.0, 12.0) | 17.0 (17.0, 19.0) | 0.043 |
|  | Female | 12.0 (10.0, 14.0) | 17.0 (15.0, 18.0) | <0.001 |
| Program | Nursing | 11.0 (8.0, 12.0) | 17.0 (15.0, 18.0) | <0.001 |
|  | Midwifery | 14.0 (13.0, 16.0) | 17.0 (16.0, 19.0) | 0.004 |
| Semester of study | Semester one | 11.0 (4.0, 15.0) | 17.0 (17.0, 17.0) | 0.109 |
|  | Semester two | 10.0 (8.0, 12.0) | 15.5 (14.0, 17.5) | 0.011 |
|  | Semester three | 12.0 (10.0, 14.0) | 17.0 (15.0, 19.0) | <0.001 |

### Participants’ ability to break bad news to patients/caretakers

The ability to break bad news to the patient/caretaker scores improved from the baseline median score of 9.0 (IQR: 7.0, 12.0) to 16.0 (14.0, 18.0) after the intervention (P value <0.001). The ability to break bad news to the patient/caretaker scores improved in all categories of sex, program and semester of study (P values <0.05) apart from participants who were in the first semester of study (P value 0.109). The findings are presented in Table 6.

**Table 6:**
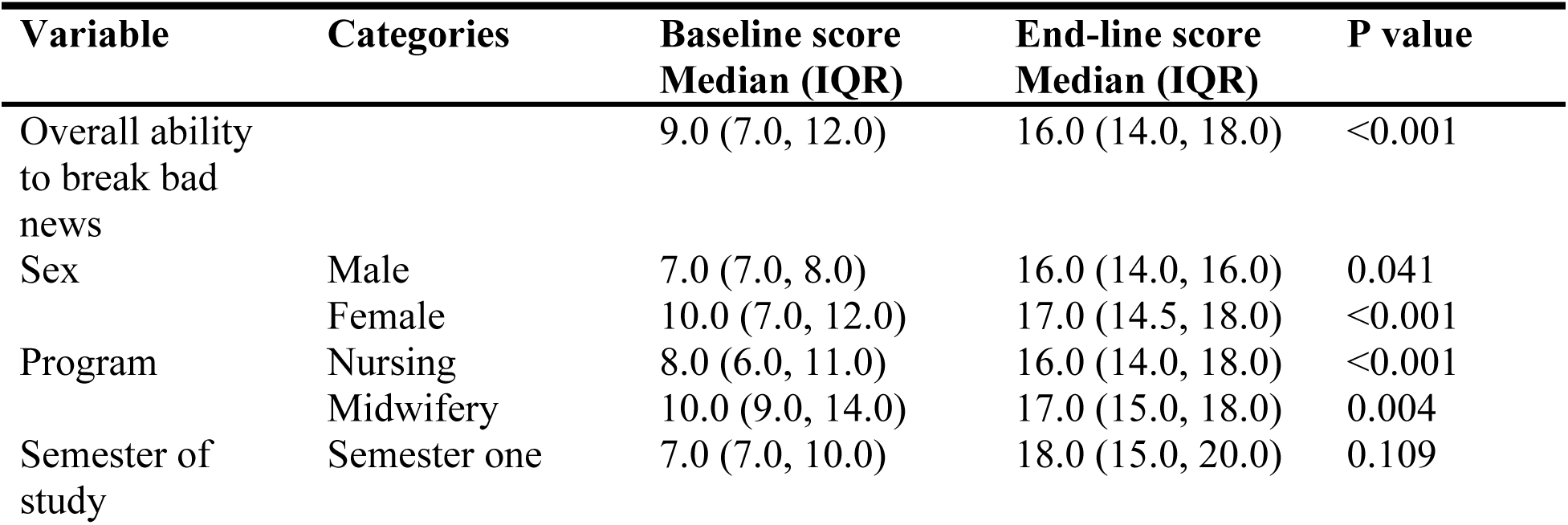
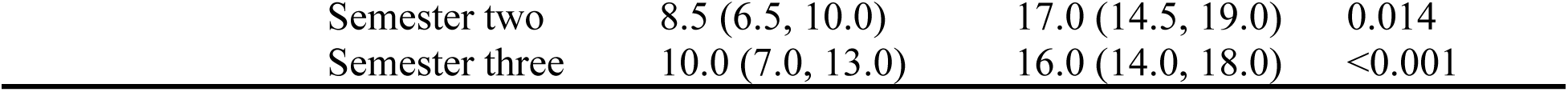
Participants’ scores of their ability to break bad news to patient/caretaker before and after the educational intervention.

## Discussion

The purpose of this study was to implement and evaluate the preliminary effectiveness of a Therapeutic communication interventional module among DNE students. Generally the study has discovered that using a targeted educational intervention can improve therapeutic communication skills of nurse trainees. The competencies addressed in this study included foundational concepts of therapeutic communication that are culturally sensitive and support the formation of effective nurse–patient relationships (8, 19–20). Through interpersonal therapeutic interactions, nurses can better meet patients’ needs and fulfill their professional responsibilities, as highlighted in Hildegard Peplau’s Theory of Interpersonal Relations (4, 21). Ethical and legal considerations were emphasized, particularly the importance of adhering to ethical principles amid evolving trends and challenges in therapeutic communication (22). The impact of the therapeutic environment on the success of nurse–patient interactions was also taught (17, 18). In emergency or crisis situations, effective communication is essential for patient safety, reducing anxiety, and ensuring clear instructions, with nurses expected to demonstrate calm, leadership, and resilience (19). The module also covered expectations, benefits, and challenges of therapeutic communication in specific contexts, including the Emergency Department (19–21), labor and delivery (22–28), care for aggressive or violent patients (29–30), and palliative care (31–34).

The use of developed courses as an intervention to improve TC can only be made more successful when combined with educational pedagogies like incorporating active teaching strategies and the use of technology in learning since most of the students prefer self-directed, immediate, exciting and immersive experiences (13). This module was delivered using blended learning and active teaching strategies including using technology to make these techniques a part of their professional lives (29, 9). Applying the principles of constructivist theory, students were encouraged to draw on their previous experiences to improve their communication skills. In the classroom, theoretical components focused on building students’ knowledge and understanding of therapeutic communication (TC). To provide a safe environment for skill development before entering clinical practice (9), students participated in simulations, demonstrations, and return demonstrations in the clinical skills laboratory. During clinical sessions, students observed TC in practice through role models. Studies indicate that both nurses and educators serve as influential role models, significantly shaping students’ learning and professional behavior in clinical settings (34, 37–38).The principal investigator (PI) actively participated in clinical learning to help students connect theoretical knowledge with real-world practice (32). Clinical sessions aimed to enhance students’ competencies and readiness for professional practice (32), bridging the gap between classroom learning and actual patient care. Although students were not formally assessed on real-time performance, they were required to maintain reflective journals to document their clinical experiences and learning (35).

### Evaluation of the TC module

This study confirmed the multifaceted effects of implementing a TC intervention on the knowledge, perceived ability and practice of TCC by nursing students. Nurses’ knowledge improved from 4 to 8.0, and their perceived confidence in practicing therapeutic communication improved from 144.0 to 164.0. The nurses’ ability to initiate a nurse-patient relationship improved from 12.0 to 17.0 while their ability to break bad news to the patient/caretaker improved from 9.0 to 16.0 after the intervention. The improved TC competency results of nursing students in this study are considered a positive effect of the TC intervention. This intervention is based on ADDIE Model and the constructivist theory that posits that students use their previous experiences to modify or create new ones. Evaluation, the last phase of the ADDIE model, is multidimensional and continuous throughout the training process as it aims to measure the extent to which the set objectives have been achieved, the competences acquired and to improve future training(15).

A previous systematic review reported preliminary evidence that interventions can effectively train nursing students in patient-centered communication (37). Studies have shown that employing multiple strategies enhances communication skills essential for establishing therapeutic nurse–client relationships (38). Nurse-led interventions using SBAR tools have also improved nurses’ knowledge and application of the Situation, Background, Assessment, and Recommendation framework, helping to reduce communication errors (3). Therapeutic communication interventions, including mirroring interviews and shared experiences, have been found to strengthen nursing students’ patient-centered communication skills (2).

This study utilized simulation, which previous research has demonstrated to significantly enhance both knowledge and practice of therapeutic communication among nursing students (39). Integrating simulation into nursing curricula is therefore recommended to improve communication skills and clinical competence. Additionally, short workshops have proven effective for refining nurses’ communication abilities, highlighting the value of structured training sessions and practical guides in enhancing communication skills for both nurses and patients (6).

### Strengths and Limitations

This study demonstrated that the therapeutic communication (TC) module effectively improved student nurses’ knowledge, perceived confidence in practicing TC, and practical TC skills. The module was implemented across classroom, clinical skills laboratory, and clinical settings using a quasi-experimental pre–post design, a robust approach that has been widely utilized in trying out educational interventions like it was in this study. The pre–post questionnaire used to assess outcomes had previously been validated for DNE students in Uganda (Cronbach’s α > 0.96). A 45-hour TC module was developed and validated through Nominal Group and Delphi techniques, incorporating input from healthcare service users, students, trainers, and regulators. This rigorous quasi-experimental approach enabled comparison of pre- and post-intervention outcomes in a real-world educational setting, enhancing the study’s external validity. The study represents the final phase of the ADDIE model framework. Limitations of the study include census sampling from a single nurse training school that, limits the generalizability of the findings to other training schools. Secondly the study focused only on the preliminary effectiveness thus the short time for posttest restricts the ability to assess the long-term retention of communication skills and their transfer to clinical practice.

### Recommendations

Longitudinal studies are needed to assess the sustained effectiveness of the TC interventional module. Additionally, research exploring patients’ or clients’ perspectives on the therapeutic communication skills demonstrated by nursing students is recommended. Finally, it is suggested that training institutions adopt and adapt this module to enhance the communication skills of future nurses, ensuring its seamless integration into nursing curricula.

## Conclusion

This study provides preliminary evidence that the educational intervention effectively enhanced nursing students’ acquisition of therapeutic communication skills. While the module significantly improved TC competence, the study did not evaluate the effectiveness of the specific teaching methods used. It is anticipated that training institutions can adopt and adapt this module to strengthen the communication skills of future nurses.

## Author’s contributions

Conceptualization of the study was done by the first author (HM). All authors (HM, AGM, RCN, and PM) did the final analysis of the study. Project administration was done by the first and second authors (HM and AGM). H.M. organized and facilitated the training. Overseeing the process of data collection was done by the first author (HM). Data analysis and interpretation were done by all authors (HM, PM, RCN and AGM). The original draft was done by the first author (HM). All authors (HM, AGM, RCN, and PM) reviewed, edited, and approved the manuscript.

“Corresponding author: Helen Mukakarisa”

## Institutional Review Board Statement

The study was conducted in accordance with the Declaration of Helsinki.

## Informed Consent Statement

Informed consent was obtained from all subjects involved in the study.

## Data Availability Statement

The raw data supporting the conclusions of this article will be made available by the authors on request.

## Acknowledgments

A special thanks to all the assessors and students who participated in the study. We acknowledge the Uganda Nurses and Midwives Examinations Board (UNMEB) for the logistical support.

## Supporting information

**S1 Table. TC interventional module content and key competencies**.

